# Dramatic Rise of Seroprevalence Rates of SARS-CoV-2 Antibodies among Healthy Blood Donors: The evolution of a Pandemic

**DOI:** 10.1101/2021.03.02.21252448

**Authors:** Maher A. Sughayer, Asem Mansour, Abeer Al Nuirat, Lina Souan, Mohammad Ghanem, Mahmoud Siag, Sallam Alhassoon

**Affiliations:** Department of Pathology and Laboratory Medicine, King Hussein Cancer Center, Amman, Jordan; Department of Radiology, and CEO, King Hussein Cancer Center

## Abstract

**Background:** The coronavirus disease 2019 (COVID-19) pandemic has resulted in more than 106 million cases of confirmed infection and more than 2.3 million deaths worldwide as of February 11th 2021. Seroprevalence studies are extremely useful in studying and assessing the epidemiological status in the community and the degree of spread. They help decision makers in implementing or relaxing mitigating measures to contain the disease in addition to other benefits.

**Objective:** To study the seroprevalence rates of SARS-CoV-2 antibodies among healthy blood donors in Jordan, at various points of time as the pandemic evolves in the community.

**Methods:** A total of 1374 blood donor were tested for the SARS-CoV-2 antibodies in 3 groups.

The first group of 746 and the second of 348 individuals were tested in June and September of 2020 respectively. The 3^rd^ group of 292 were tested in early February of 2021. We utilized a qualitative assay that uses Electrochemiluminescence method (ECLIA) that has a specificity and sensitivity of 99.8% and 100% respectively.

**Results:** The first 2 groups representing the months of January to September of 2020, where the number of confirmed Covid-19 cases were several hundred to 3000 showed a seroprevalence rate of 0% (95% CI 0.00%, 0.51%). The 3^rd^ group representing late January and early February 2021 when the number of reported confirmed case has reached 100 folds the numbers of September 2020, showed a seroprevalence of 27.4% (95% CI 22.5% and 32.9%).

**Conclusions:** a dramatic rise in seroprevalence of SARS-CoV-2 antibodies was seen among healthy blood donors in Jordan in parallel with wide-spread intracommunity transmission of the disease. This information is useful to assess the degree of herd immunity and provides for better understanding of the pandemic.

## Introduction

The coronavirus disease 2019 (COVID-19) pandemic has resulted in more than 106 million cases of confirmed infection and more than 2.3 million deaths worldwide as of February 11th 2021 (European Centre for Disease Prevention and Control, 2021).

Population based Seroprevalence studies are extremely important to understand the evolution of the pandemic and to estimate infection rates and prevalence. They are important for calculating absolute risks of the infection as well as death rates and to predict the spread of the virus in communities based on the level of herd immunity after infections and or vaccination. They are also important for planning and monitoring the impact of implementation and relaxation of epidemic mitigation policies (Busch and Stone, 2021)

The true prevalence of the infection is believed to be several times more than the number of PCR confirmed cases because of the large number of asymptomatic infections and or mild infections that went untested especially early in the pandemic (Huang et al., 2020, McLaughlin et al., 2020, Busch and Stone, 2021. The ratio of estimated to reported infections can range up 12.5 (Bajema et al., 2020)

Numerous population seroprevalence studies were conducted (Bajema et al., 2020, Lai et al., 2020, Chughtai et al., 2020, McLaughlin et al., 2020, Naranbhai et al., 2020, Sam et al., 2021, Sutton et al., 2020, Menachemi et al., 2020, Silveira et al., 2020, Kar et al., 2021, Vena et al., 2020, Pollán et al., 2020, Bogogiannidou et al., 2020, Poustchi et al., 2020., Shields et al., 2020, Ng et al., 2020, Figueiredo-Campos et al., 2020, Stringhini et al., 2020, Havers et al., 2020, Ho et al., 2020, Xu et al., 2020, Qutob et al., 2020, Capai et al., 2020, Sood et al., 2020, Godbout et al., 2020, Rostami et al., 2020) in efforts to estimate the true prevalence of the COVID-19 infection. The largest seroprevalence study was the one by Bajema et al. from the USA which showed by September 2020 that the estimated population seroprevalence to be less than 10% in the majority of tested communities, although it ranged from 0 up to 23.3% in the highest hit areas. Obviously the wide variation reflects the level of transmission in the tested communities and number of PCR confirmed reported cases. (Bajema et al., 2020).

Those population-based seroprevalence studies varied in the targeted populations tested and the recruitment strategies used which may explain some of the variation. Among the populations tested are healthy blood donors (Sughayer et al., 2020, Slot et al., 2020, Daniel J. Nesbitt et al., 2021, Gallian et al., 2020, Olariu et al., 2021, Younas et al., 2020, Banjar et al., 2021, Uyoga et al., 2021, Busch and Stone, 2021, Martinez-Acuña et al., 2020, Saeed et al., 2021, Slot et al., 2020, Fiore et al., 2021). Again the seroprevalence among healthy blood donors and other population groups studied varied according to the community tested and the time of testing in terms of the pandemic evolution (Lai et al., 2020, Rostami et al., 2020)

In this longitudinal study we aimed to estimate the seroprevalence of Covid-19 among healthy blood donors in Jordan at different points in time to assess the degree of community spread and herd immunity and to further understand the evolution of the pandemic. This is the first longitudinal seroprevalence study in blood donors and the first from Jordan.

## Methods

### Subjects and samples

Left over sera and or plasma collected routinely during the process of blood or apheresis platelet donations were used for the study. The donors were healthy asymptomatic subjects between the ages of 18 and 63 who underwent routine screening to determine their acceptability for donation as per standard practice. The sera were tested in 3 batches at 3 different times. The first batch consisting of 746 samples representing the period of January to June 2020 was tested in June of that year. The second batch of 348 samples were collected and tested in September of 2020 while the third batch of 292 samples were collected during the period of January 28-Februay 5, 2021 and tested in the same period. Thus the total number of donors tested in the study was 1374.

## Testing Methodology

The samples were tested according to the manufactures recommendations using a commercially available FDA approved kit for total immunoglobulins against SARS-Cov-2. The test was performed on Cobas 6000 or Cobas Pro, Roche analyzers, using the Elecsys-Anti-SARS-CoV-2 kit (Roche Diagnostics GmbH, Mannheim). This test is a qualitative assay that uses Electrochemiluminescence Method (ECLIA) which is an immunoassay for the in vitro qualitative detection of antibodies (including IgG) to SARS-CoV-2 in human serum and plasma. The assay uses a recombinant protein representing the nucleocapsid (N) antigen for the determination of antibodies against SARS-CoV-2. The test was validated by the manufacturer using 5272 samples including blood donors, diagnostic routine, other corona viruses and common cold panels. The specificity was determined to be 99.8% (95% CI, 99.7%-99.9%). while the sensitivity was 100% (95% CI, 88.3%-100.0%).

In house validation using serum samples obtained from previously RT-PCR-confirmed Covid-19 infected patients who have recovered at least one month prior to sampling was performed.

## Statistics

Descriptive statistics were used to analyze the results. Chi square statistics were used to compare the seropositive donors’ characteristics versus the seronegative ones. We adjusted the estimated prevalence to test’s sensitivity and specificity using an online tool: http://www2.univet.hu/users/jreiczig/CI4prevSeSp/calc02/index.php as. per methods described by Lang and Reiczjkl (Lang and Reiczigel, 2014).

### Ethical approval

All necessary approvals were obtained.

## Results

The demographics of the donors and the period of donation are summarized in table 1. The donors were mostly males (86%) and from the capitol city of Amman, Jordan (78%).

**Table 1.**

| Demographics of the 1374 Healthy Donors- No. (%) |  |
| --- | --- |
| Male | 1177(85.7%) |
| Female | 197 (14.3%) |
| Residential area: Amman | 1077 (78.4%) |
| West Amman | 706 (65.6%) |
| East Amman | 371 (34.4%) |
| District outside Amman | 297 (21.6%) |
| North | 128 (9.3%) |
| Center | 138 (10.1%) |
| South | 31 (2.3%) |
| Period |  |
| January-2020 | 104 (7.6%) |
| February-2020 | 89 (6.5%) |
| March-2020 | 90 (6.6%) |
| April-2020 | 147 (10.7%) |
| May-2020 | 229 (16.7%) |
| June-2020 | 75 (5.4%) |
| Septemper-2020 | 348 (25.3%) |
| 28 Jan- Feb 5<sup>th</sup>, 2021 | 292 (21.3%) |

The first group was previously reported in a preprint (Sughayer et al., 2020) and revealed along with the second group a seroprevalence rate of 0% in each group (95% CI 0.00%, 0.51%).

The third group on the other hand which represents the most recent period of January 28-February 5, 2021, showed a positive serological test for the SARS CoV-2 in 80 of 292 donors; a crude seroprevalence rate of 27.4%. The adjusted estimated seroprevalence rate is 27.3% (95% confidence intervals 22.5% and 32.9%) The demographics and characteristics of the seropositive donors in comparison to the seronegative ones are shown in Table 2. Most of those who tested positive (85%) were in the age group of 18-40 years. However there was no statistically significant difference between the seropositive and negative donors in terms of gender, age, blood group or residence. Males and females were almost equally affected (27.6% vs 26.3%).

**Table 2.** Comparison of seronegative and seropositive donors

Comparison of seronegative and seropositive donors
| Category |  | Seropositive donors<br>80 of 292 | Seronegative donors<br>212 of 292 | p-value | Crud Prevalence rate |
| --- | --- | --- | --- | --- | --- |
| number of donors |  | 80 (%) | 212 (%) |  | 27.4% |
| Male |  | 70 (87.5%) | 184 (86.8%) | 0.9 | 27.6% |
| Female |  | 10 (12.5%) | 28 (11.8%) |  | 26.3% |
| Age (yr) | 18-30 | 47 (58.8%) | 116 (54.7%) | 0.4 | 28.8% |
|  | 31-40 | 21 (26.3%) | 52 (24.5%) |  | 28.8% |
|  | 41-50 | 11 (13.7%) | 24 (11.3%) |  | 31.4% |
|  | 51-65 | 1 (1.2%) | 11 (5.2%) |  | 8.3% |
|  | Unknown | 0 | 9 (4.2%) |  |  |
| Blood group: | O | 33 (41.3%) | 88 (41.5%) | 0.5 | 27.3% |
|  | A | 23 (28.8%) | 73 (34.4%) |  | 24.0% |
|  | B | 15 (18.7%) | 37 (17.5%) |  | 28.8% |
|  | AB | 9 (11.2%) | 14 (6.6%) |  | 39.1% |
| Residential Location | North | 7 (8.8%) | 33 (15.6%) | 0.3 | 17.5% |
|  | Center including Amman | 68 (85.0%) | 168 (79.2%) |  | 28.8% |
|  | South | 5 (6.2%) | 11 (5.2%) |  | 31.3% |
| Covid-19 | PCR-Confirmed past infection | 16 (20%) | 0 |  |  |
|  | No/negative PCR | 47 (58.8%) | 180 (84.9%) |  |  |
|  | No information available | 17 (21.2%) | 32 (15.1%) |  |  |

One fifth of the seropositive donors were retrospectively found to have been confirmed positive for the covid-19 infection by the PCR test. Forty seven (58.8%) were not known to have the disease and so did not undergo or underwent a PCR test with negative result. There was no information with regards to previous infection for 17 of the seropositive donors.

## Discussion

The importance of serological testing for the SARS COV-2 antibodies have been previously highlighted (Busch and Stone, 2021, Raoult, 2021). Among the advantages of such testing is the understanding of the evolution of the pandemic in terms of having a rough estimation of the prevalence of the infection. This will serve health planners and decision makers to properly enforce or relax mitigation measures. And most importantly at these times as vaccines are being rolled out it serves in estimating the risk rates for the infection, the degree of herd immunity and helps prioritizing vaccine recipients.

In this study we measured the seroprevalence rates in healthy blood donors at three points in time. The results are striking in that they show a dramatic change from 0% early and in the middle of the pandemic, up to 27.4% in February of 2021. These findings would be reasonable if we consider the cumulative number of confirmed cases around these times in Jordan. Figure 1 shows the cumulative daily cases of Covid-19 in Jordan. In the first period up till June 2020 there were only several hundred confirmed cases which increased gradually to around 3000 cases in early September (WHO, 2021). However towards the end of September and afterwards the cumulative number of cases started a steep rise so that on Februay 5, 2021 the cumulative number of confirmed cases was more than 100 folds that of September 2020. It is worth mentioning that a strict lockdown was in effect till early June that was gradually relaxed over three months with full opening of all sectors including schools and international travel in September 2020. It is clear that the first wave of the Covid-19 pandemic in Jordan has actually started in late September when the community was fully open and the intracommunity spread became evident. Before that the several hundred cases were actually limited to transmission within known specific hot foci. This explains the extremely low seroprevalence initially found in June and September of last year, as the infection transmission was under strict control with quarantine routinely imposed on all contacts of index cases.

**Figure 1.**
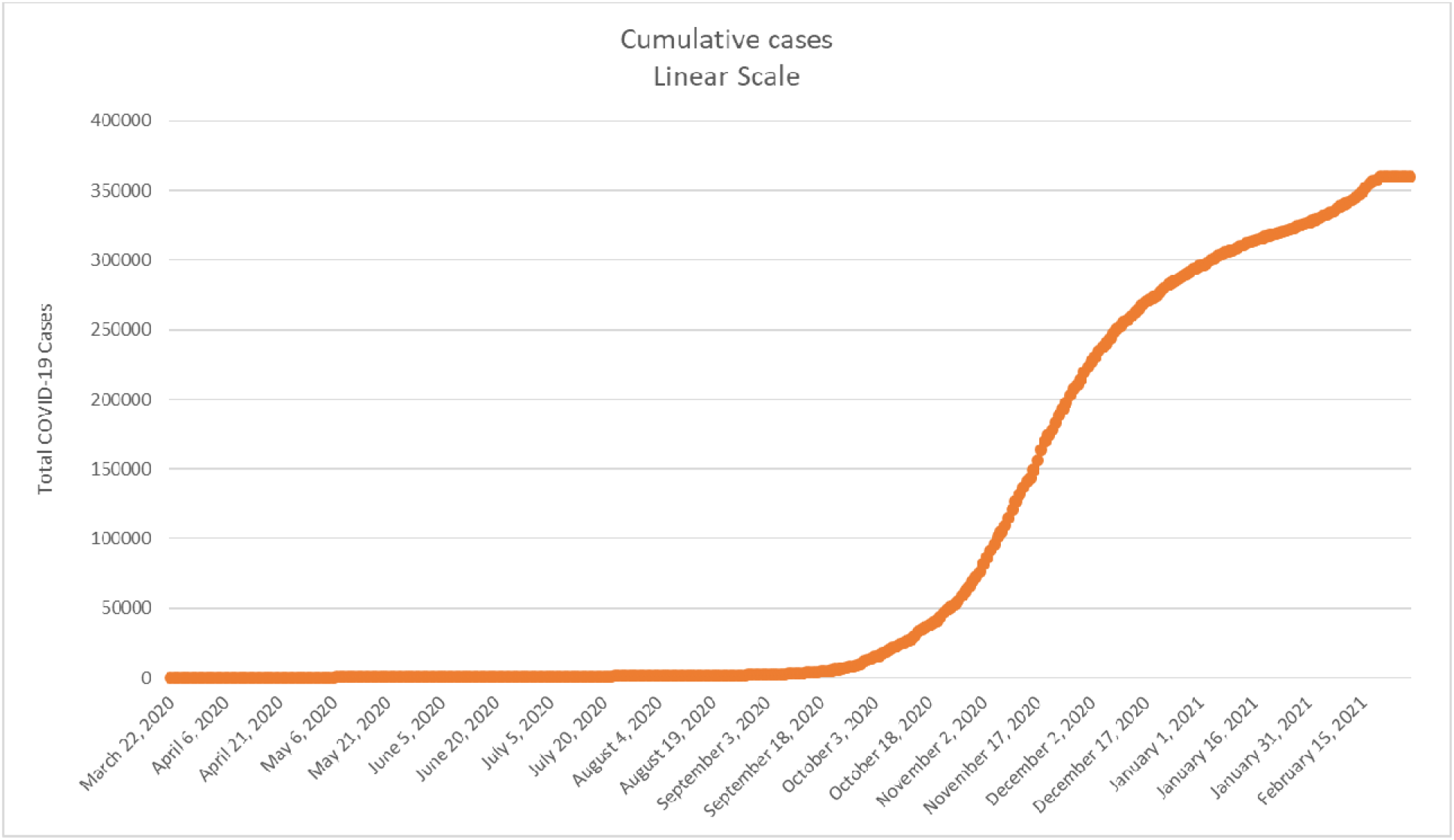

The current (early February 2021) crude seroprevalence rate of 27.4%, if can be generalized to the population at large would mean that the number of cases is roughly 2.7 million in a population of 10 million in Jordan. This, if true, means that there are 8 cases for every confirmed case. This high ratio of estimated to reported cases is similar to some of the highest ratios reported by Bajema et al. in the USA (Bajema et al., 2020). Looking at it from a different angle we can also see that in our rather small sample one fifth of the donors were previously confirmed positive by the PCR test so we may assume a ratio of estimated to reported cases to be 5 at the lowest estimate.

Most of our seropositive donors (85%) were young; below 40 years of age. This finding helps in setting the vaccination priority for those older than 40 years. Other studies have shown inconsistent findings with either similar age group distribution to our cohort or the opposite with higher rate in older individuals (Bajema et al., 2020, Martinez-Acuña et al., 2020, Vena et al., 2020, Olariu et al., 2021).

Our findings are also in line with previous studies which showed that early in the pandemic the seroprevalence of Covid-19 in blood donors or other nontargeted populations was very low ranging from 0% to 2.0% (Erikstrup et al., 2021, Godbout et al., 2020, Qutob et al., 2020, Xu et al., 2020, Ho et al., 2020, Banjar et al., 2021, Nesbitt et al., 2021Slot et al., 2020, Fiore et al., 2021, Saeed et al., 2021). However other studies especially those from hardly hit communities early in the pandemic showed relatively high rates up to 23% (Percivalle et al., 2020).

Our current high seroprevalence rate (as of early February 2021) is similar to that of communities that were hardly affected by the pandemic such as that of New York City and Chelsea, Massachusetts (Bajema et al., 2020, Naranbhai et al., 2020).

With regards to blood group association with decreased risk for Covid-19 infection in blood group O (Gallian et al., 2020) our limited data did not reveal such an association.

Our study to our knowledge is the first longitudinal study of covid-19 seroprevalence in healthy blood donors.

There are some limitations in our study including the small number of the blood donors in the 3rd group. In addition the blood donors are mostly males and of younger age groups. However the age groups represented in our study constitute around 60% of the Jordanian population, the rest being mostly children under the age of 18. Thus it may be difficult to draw generalization as there may be selection bias and nonrepresentation of the entire population. Further studies are recommended to include children and the elderly.

## Data Availability

No data available

